# Room-Specialized Mixture-of-Experts for In-Home ADL Recognition with Ambient Sensors

**DOI:** 10.64898/2026.06.10.26355390

**Authors:** Venkatanand R. Addepalli, Praveen Rao, Andrew Kiselica, Erich Kummerfeld, Nader Abdalnabi, Knoo Lee

## Abstract

Monitoring activities of daily living (ADLs) in the home is a promising approach for tracking dementia progression in older adults. While ambient sensor-based ADL systems are well-studied, most existing ADL recognition systems rely on globally trained models that ignore the spatial organization of in-home activities. In real deployments, where training data are sparse and highly home-specific, global transformer models may fail to capture room-dependent behavioral structure. We propose a deterministic Mixture of Experts (MoE) architecture for in-home ADL recognition, in which each expert is a compact transformer specialized to one room of the home (bedroom, kitchen, bathroom, living area). Input segments are routed using a deterministic gating strategy based on room-level motion activity and time-of-day priors for sleep-related behaviors. Unlike learned routing networks, the proposed gate encodes domain knowledge about where ADLs are likely to occur, reducing model complexity under limited per-home training data. By decomposing ADL recognition into room-specific activity spaces, the proposed architecture reduces competition between dominant and low-frequency activities under highly imbalanced residential data. We evaluated the system on data collected via low-cost ambient sensors (motion, light, temperature, humidity) and Raspberry Pi edge devices across five homes, with ground-truth ADL labels provided by participants and caregivers. Across the five homes, the proposed MoE consistently outperformed global transformer, 1D CNN, and Random Forest baselines, achieving macro-F1 scores ranging from 0.60 to 0.88, highlighting the importance of home-specific modeling in real-world deployments. These findings suggest that room-aware expert specialization may provide a practical and interpretable strategy for low-data ADL recognition in real-world residential environments.

## I. Introduction

Alzheimer’s disease and related dementias (ADRD) are increasingly prevalent among older adults and are associated with progressive decline in activities of daily living (ADLs), including bathing, meal preparation, medication management, and mobility within the home. Because functional decline in ADLs is a defining characteristic of cognitive impairment progression, continuous in-home monitoring has emerged as a promising strategy for longitudinal assessment in aging populations[1].

Prior work has established several influential ambient sensing datasets for ADL recognition, including CASAS, CASD, and ARAS [2-5]. These datasets have supported a wide range of activity recognition approaches using classical machine learning and, more recently, deep learning methods [6,7]. However, many existing approaches treat ADL recognition as a uniform multi-class prediction problem using globally trained models, despite the strong spatial organization of activities within residential environments.

Public benchmark datasets are typically collected in controlled smart-home environments with standardized activity durations and scripted routines. In contrast, real-world ADLs are highly variable, interrupted, overlapping, and strongly class-imbalanced. Frequent behaviors such as sleep dominate daily activity streams, whereas clinically important activities such as medication intake may occur only rarely. Standard balancing approaches, including oversampling and undersampling methods[8, 9], often distort the temporal structure of sequential sensor data and may reduce deployment realism.

In addition, many prior approaches rely on globally trained architectures originally designed for large benchmark datasets[10]. Real residential deployments instead operate under limited per-home training data, substantial between-home behavioral heterogeneity, and constrained edge-computing environments. Under these conditions, large monolithic models may be poorly matched to the spatial and temporal structure of in-home ADLs.

To address these challenges, we propose a deterministic-gated Mixture-of-Experts (MoE) architecture in which compact transformer experts specialize to room-specific activity patterns [11]. Rather than learning routing behavior from limited training data, the proposed gating mechanism incorporates domain knowledge about where ADLs occur within the home. This design reduces model complexity while preserving interpretable spatial structure in residential activity streams. To our knowledge, this is the first application of mixture-of-experts modeling for per-home ADL recognition using ambient sensing data.

## II. Methods and Materials

### A. Study Design and Participants

We recruited nine community-dwelling older adults with mild dementia. Each participant was paired with a co-resident caregiver, and continuous ambient sensing was deployed within participants’ homes. For supervised ADL labeling, caregivers retrospectively logged participant activities over an initial one-month annotation period using a structured reporting template and standardized study guidelines at predefined times each day to improve recall consistency. To capture representative weekday and weekend behavioral variation while minimizing routine-related variance, activity logging was performed on two weekdays and one weekend day per week. Logging focused on a predefined set of 13 primary ADLs derived from IADL and Katz ADL frameworks, with optional sub-activity annotations when additional detail was available [12]. Of the nine enrolled homes, five produced usable ADL datasets for downstream analysis, while four were excluded because of insufficient annotation quality or incomplete activity records.

### B. Smart Home Configuration

Each home was instrumented with four passive infrared (PIR) motion sensors, two door sensors, and one bathroom multi-sensor measuring light, temperature, humidity, and motion. A Raspberry Pi 4 edge device was configured to record sensor events locally and transmit data securely to a cloud-based storage and analysis platform. The complete sensor layout is shown in Fig. 1.

**Fig. 1.**
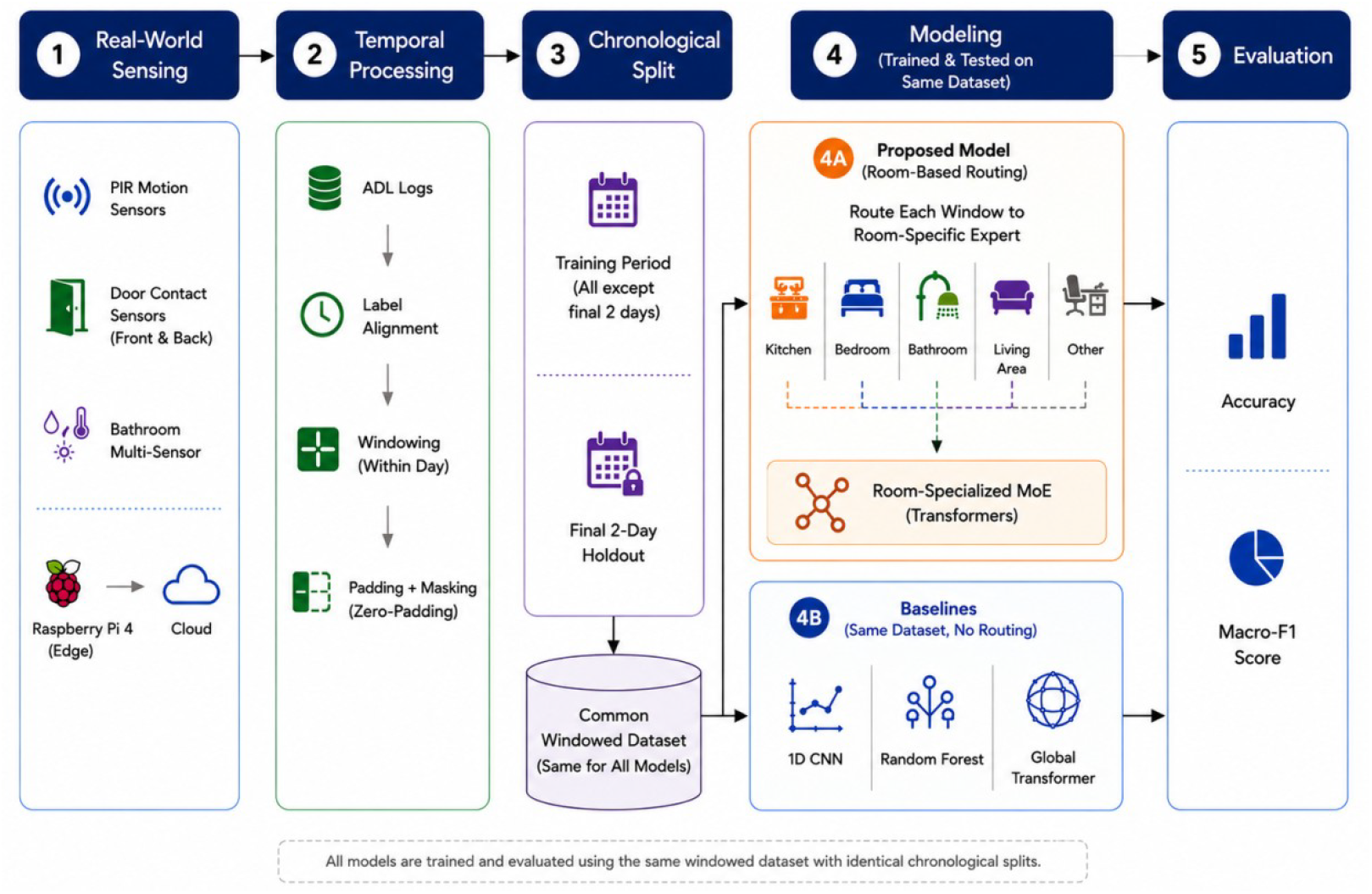
Overall workflow of the process.

### C. Data Preprocessing

The collected data comprises two raw streams:

(i) sensor events recorded by the smart home infrastructure,
(ii) ADL labels manually coded by the dyad. Each ADL record is a tuple of the form (*Time_Start,_Time_End*, Actor, Activity). To unify the two streams, we resampled both to a one-minute frequency.

For the ADL stream, when n activities were reported within a single labeled interval, we partitioned the interval into n equal sub-windows:

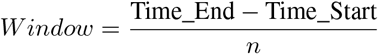

and assigned the reported activities sequentially to each subwindow. This enforces our working assumption of one activity per minute and yields a uniform temporal grid. The sensor stream contains a mix of discrete and continuous variables. *Motion_State* events are inherently discrete (presence or absence of motion at a given timestamp), whereas light and temperature are continuous quantities reported only when the sensor detects a change. Following our prior pilot study, we applied [cubic spline] interpolation between successive observations to obtain a continuous, minute-resolution signal for each continuous channel[13]. The resampled channels were then inner-joined on timestamp to produce a single, unified sensor table.

### D. Room-Based Segmentation

Exploratory analysis of the labeled activity distributions demonstrated strong location dependence among ADLs. Most activities are confined to a single room: cooking, eating, dishwashing, and medication management occur primarily in the kitchen; sleeping and dressing in the bedroom; hygiene and toileting in the bathroom; and leisure, napping, and some eating in shared living spaces. A second class of activities, however, is not confined to a single room but instead spans multiple locations or is characterized by movement between them. Mobility and Transportation, for example, are better captured by door-traversal events than by the motion signature of any one room.

Based on these patterns, the dataset was restructured into location-specific temporal segments and routed to one of two kinds of experts. Single-room activities are assigned to room experts, each operating on the sensor channels of one room. Movement-based activities are assigned to span experts that operate on door-sensor channels spanning multiple rooms. This decomposition enables location-conditioned activity modeling and reduces ambiguity between participant and caregiver activity patterns within shared residential environments.

A *segment* is defined as a contiguous interval of constant activity label within a single calendar day. Segment boundaries are introduced at activity transitions and at midnight to prevent overnight sleep intervals from spanning the train/test split.

Each segment is divided into non-overlapping windows of length *L* minutes. A segment of duration *D* minutes yields ⌈*D/L*⌉ windows. The final window of each segment is zero-padded to length *L*, and an associated binary length mask **m** ∈{ 0, 1} ^*L*^ marks real (*m*_*t*_ = 1) versus padded (*m*_*t*_ = 0) positions.

Padded positions are excluded from both attention computation and temporal pooling so that they contribute no signal to the model prediction.

We evaluate on

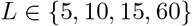

minutes to examine the effect of temporal context granularity on ADL recognition performance, ranging from short-duration activity transitions to longer sustained behavioral routines.

### E. Class Imbalance Handling

Activity durations are highly imbalanced in real-world data. In our corpus, sleep typically accounts for 6-10 hours per day while activities such as dressing or medication management may last only a few minutes. Rather than applying synthetic oversampling, which fabricates sensor co-occurrence patterns absent from real data, or undersampling, which discards informative segments, we address imbalance through two complementary mechanisms. First, the segmentation procedure of previous section produces multiple windows per long segment, which naturally increases the effective number of training examples for long-duration activities without distorting their sensor signatures. Second, the cross-entropy loss for each expert uses class-balanced weights computed from the trainingset class distribution, clipped to prevent extreme reweighting on very rare classes. No segments are truncated, discarded, or synthesized at any point in the pipeline.

### F. Model Building

The proposed architecture is a Mixture of Experts (MoE) in which each expert is a compact transformer encoder specialized to a single room of the home, and a deterministic gating function assigns each input segment to exactly one expert [14].

*Per-room expert*. Each expert *e* ∈ {1, …, *E* } has its own input sensor set *S*_*e*_ (motion, temperature, light, and optionally humidity) and active class set *C*_*e*_ with *K*_*e*_ = |*C*_*e*_| classes. The input window is 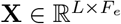, where *F*_*e*_ = |*S*_*e*_| + 3. The three additional channels are metadata appended to every timestep: the segment duration and a cyclic encoding of the hour of day, sin(2*πh/*24) and cos(2*πh/*24), where *h* is the fractional hour (e.g., 14.00 for 14:00, 14.03 for 14:02) taken from the `DateTime` column. The cyclic encoding preserves the periodicity of daily rhythms, midnight and 23:59 are adjacent rather than 23 hours apart, while the fractional hour preserves minute-level resolution.

Each timestep is projected into a *d*-dimensional latent space:

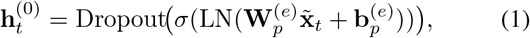

Where 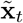 is the standardized input at timestep *t, σ* is the GELU activation, 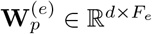 is the projection matrix, and 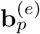 its bias. Stacking over *t* gives **H**^(0)^ ∈ R^*L×d*^, to which a learned positional embedding **P**^(*e*)^ ∈ R^*L×d*^ is added. A single encoder layer with two attention heads applies masked multi-head selfattention (MHSA) followed by a position-wise feed-forward block (FFN), each with a residual connection and LayerNorm:

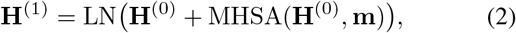

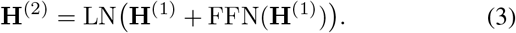

The length mask **m** ∈ { 0, 1} ^*L*^ ensures that no valid position attends to a padded position in the attention softmax. temporal dimension is collapsed by masked mean pooling over valid timesteps:

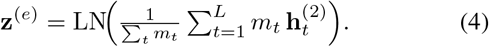

The pooled vector is mapped to class logits through a twolayer head:

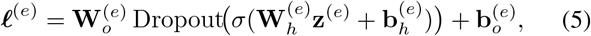

where 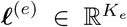. By construction, each expert produces logits only over its own class set *C*_*e*_.

*Rule-based gating*. The gate *g* deterministically assigns each segment to exactly one expert using domain knowledge:

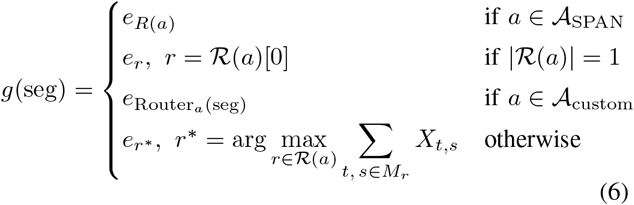

where *a* is the activity label, ℛ(*a*) is the set of candidate rooms for *a*, and *M*_*r*_ is the set of motion sensors in room *r*. The set 𝒜_SPAN_ contains multi-room activities such as Mobility and Transportation, routed to predefined door-based span experts. For sleep-related activities (𝒜_custom_), the router Router_*a*_ applies a temporal heuristic: the bedroom is selected if the segment starts within the night interval (21:00–09:00), otherwise the living area. Remaining multi-room activities are routed to the room with the greatest motion activity during the segment. The gate is parameter-free and relies solely on domain knowledge and sensor evidence, which is well-suited to the low-data per-home regime where a learned gating network would risk overfitting.

#### Independent expert training

Because the gate is deterministic, each expert is trained independently on the subset of training segments routed to it, and the total objective decomposes additively with no gradient flow between experts:

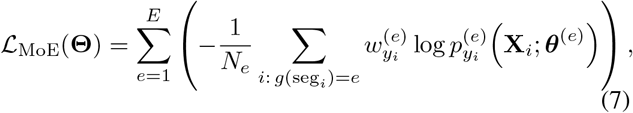

where *N*_*e*_ is the number of training segments routed to expert *e*, 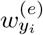 are class-balanced weights over 𝒞_*e*_, ***θ***^(*e*)^ denotes the trainable parameters of expert *e*, and 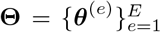 is the full parameter set.

### G. Baseline Models

We compare the proposed MoE framework against three baseline models using identical train/test splits and identical input windows.

1) **Global Transformer:** A single Transformer encoder with the same depth and attention configuration as an individual MoE expert (one encoder layer, two attention heads, and embedding dimension *d* = 16) operating over the union of all sensor channels and activity classes. The architecture follows the original Vaswani encoder design with sinusoidal positional encoding and a learned CLS token for sequence pooling.
2) **One-Dimensional Convolutional Neural Network (1D CNN)[15, 16]:** The network consists of two stacked Conv1D blocks, each defined with 32 convolutional filters per layer. The temporal dimension is collapsed using the same masked mean pooling operation as the MoE experts to ensure equivalent handling of padded positions. A final linear classification layer produces activity logits. The total model capacity is approximately 5K trainable parameters.
3) **Random Forest[15]:** Hand-crafted statistical features are extracted from each window, including per-sensor mean, standard deviation, minimum, maximum, range, and count of nonzero activations over valid timesteps. Additionally, the three metadata channels from the first valid timestep are included. The resulting feature vector is used to train a Random Forest classifier consisting of 400 trees with no maximum depth, a minimum of two samples per leaf, and balanced class weighting. Random Forest models are widely used as classical baselines for ambient ADL recognition.

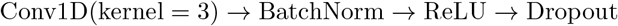

### H. Inference testing

Predictions are made per window at test time. The MoE routes each test window through its gate to a single expert, which outputs a probability distribution over its class set and set of defined hyperparameters across all the locations; the predicted activity is the argmax. The baselines output a distribution over all activity classes directly. The *No Activity* logit is masked at inference for all models, consistent with its treatment as a background class during training. Per-class precision, recall, and F1-score are computed in the standard way. The reported headline metric is macro-averaged F1-score, which weights each activity class equally regardless of support — appropriate for class-imbalanced data where rare clinically-relevant activities should not be drowned out by frequent ones.

## III. Results

We evaluated the proposed MoE, Global Transformer, 1D CNN, and Random Forest models using identical train/test splits and input windows across five homes. The proposed MoE contained approximately 2,820 trainable parameters per expert distributed across room-specialized experts, compared with approximately 15,000 parameters for the Global Transformer baseline.

Across all homes and window lengths, the proposed MoE achieved the highest macro-F1 among all evaluated models, demonstrating that room-specialized decomposition provides a consistent advantage over globally trained architectures in low-data residential sensing environments (See Table. 2). At L=5minutes, the MoE achieved macro-F1 scores ranging from 0.60 to 0.88, whereas the Global Transformer baseline remained substantially lower across most homes. These results suggest that room-specialized decomposition is substantially better matched to low-data residential deployments than globally trained transformer architectures. Training times for the 1D CNN and Random Forest baselines were approximately 3 minutes per home.

Table 3 summarizes performance across window length *L* ∈ { 5, 10, 15, 60} minutes. Increasing temporal context did not improve performance across homes. Longer windows occasionally improved recognition of sustained activites such as sleep, but often reduced sensitivity to shorter or lower-frequency ADLs including cooking and dressing.

**TABLE I.** Sample of the datasets from site01.

| DateTime | Kitchen_Motion | Kitchen_Temp | Kitchen_Luminance | Activity |
| --- | --- | --- | --- | --- |
| 2025-07-26 18:14:00 | 0 | 76.64 | 12.25 | Cooking |
| 2025-07-26 18:15:00 | 1 | 76.66 | 12.08 | Cooking |
| 2025-07-26 18:16:00 | 0 | 76.64 | 11.90 | Cooking |
| 2025-07-26 18:47:00 | 1 | 76.64 | 9 | Eating |
| 2025-07-26 18:48:00 | 1 | 76.63 | 9 | Eating |
| 2025-07-26 18:49:00 | 0 | 76.63 | 9 | Eating |

**TABLE II.** Performance (MACRO F1-SCORES) Comparison Across Different Time Windows for Location-Based Models.

| Location | 5 minutes<br>(MoE / Global / RF / CNN) | 10 minutes<br>(MoE / Global / RF / CNN) | 15 minutes<br>(MoE / Global / RF / CNN) | 60 minutes<br>(MoE / Global / RF / CNN) |
| --- | --- | --- | --- | --- |
| SITE 01 | 0.596 / 0.151 / 0.196 / 0.167 | 0.538 / 0.114/ 0.323/ 0.124 | 0.456 / 0.184/ 0.264/ 0.154 | 0.744 / 0.212 / 0.238/ 0.149 |
| SITE 02 | 0.670 / 0.189 / 0.193 / 0.230 | 0.734 / 0.217/ 0.245/ 0.209 | 0.614 / 0.276/ 0.267/ 0.228 | 0.683 / 0.297 / 0.298 / 0.179 |
| SITE 03 | 0.885 / 0.126 / 0.136 / 0.218 | 0.846 / 0.216 / 0.245/ 0.209 | 0.883 / 0.161/ 0.225/ 0.220 | 0.886 / 0.107 / 0.243 / 0.225 |
| SITE 04 | 0.679 / 0.570 / 0.448 / 0.344 | 0.900 / 0.346/ 0.139/ 0.232 | 0.917 / 0.544 / 0.092/ 0.218 | 0.677 / 0.596/ 0.098/ 0.256 |
| SITE 05 | 0.683 / 0.189 / 0.165 / 0.232 | 0.664 / 0.176 / 0.514/ 0.446 | 0.568 / 0.157/ 0.413/ 0.371 | 0.694 / 0.270 / 0.438 / 0.485 |

**TABLE III.** Per-CLASS F1 SCORES OF THE MOE AND Global Transformer models across sites and time windows.

| Activities | MoE<sup>†</sup> |  |  |  |  |  |  |  |  |  |  |  |  |  |  |
| --- | --- | --- | --- | --- | --- | --- | --- | --- | --- | --- | --- | --- | --- | --- | --- |
|  | SITE 01 |  |  | SITE 02 |  |  | SITE 03 |  |  | SITE 04 |  |  | SITE 05 |  |  |
| Cook | 0.90(9) | 0.83(5) | **1.00(1)** | 0.86(4) | 0.00(2) | **0.00(1)** | 0.59(22) | 0.54(11) | **0.40(4)** | - | - | - | - | - | - |
| Dress | 0.92(7) | 0.67(4) | **0.80(2)** | 0.80(8) | 1.00(5) | **0.80(4)** | 0.77(8) | 1.00(5) | **1.00(3)** | 1.00(9) | 0.67(6) | **0.80(3)** | - | - | - |
| Eat | 0.48(9) | 0.38(5) | **1.00(1)** | 0.00(7) | 0.36(4) | **0.00(1)** | 0.25(22) | 0.40(11) | **0.29(4)** | 1.00(9) | 1.00(42) | **1.00(5)** | - | - | - |
| Hygiene | 0.86(4) | 0.67(2) | **1.00(1)** | - | - | - | 0.64(14) | 0.77(8) | **0.80(5)** | 0.95(108) | 1.00(55) | **1.00(10)** | - | - | - |
| Leisure | 1.00(15) | 1.00(8) | **1.00(2)** | 0.93(148) | 1.00(76) | **1.00(17)** | 1.00(98) | 1.00(49) | **1.00(10)** | 0.59(128) | 0.54(65) | **0.60(14)** | 0.24(88) | 0.62(44) | **0.22(8)** |
| Sleep | 0.99(265) | 0.98(134) | **0.98(2)** | 0.92(223) | 1.00(103) | **0.95(21)** | 0.99(198) | 1.00(100) | **1.00(19)** | 0.85(202) | 0.80(101) | **0.81(19)** | 0.84(285) | 1.00(87) | **0.84(16)** |
| Toileting | - | - | - | 1.00(8) | 1.00(6) | **1.00(6)** | 0.33(3) | 0.57(2) | **0.50(2)** | 0.81(21) | 1.00(14) | **1.00(4)** | 1.00(4) | 1.00(2) | **1.00(2)** |
| Activities | Global Transformer<sup>‡</sup> |  |  |  |  |  |  |  |  |  |  |  |  |  |  |
|  | SITE 01 |  |  | SITE 02 |  |  | SITE 03 |  |  | SITE 04 |  |  | SITE 05 |  |  |
| Cook | 0.00(9) | 0.20(5) | **0.00(1)** | 0.00(4) | 0.00(2) | **0.00(1)** | 0.00(22) | 0.12(11) | **0.36(4)** | - | - | - | - | - | - |
| Dress | 0.33(7) | 0.00(4) | **0.44(2)** | 0.32(8) | 0.00(5) | **0.38(4)** | 0.00(8) | 0.00(5) | **0.00(3)** | 0.21(9) | 0.00(6) | **0.00(3)** | - | - | - |
| Eat | 0.55(9) | 0.00(5) | **0.00(1)** | 0.12(7) | 0.00(4) | **0.00(1)** | 0.35(22) | 0.00(11) | **0.53(4)** | 0.00(9) | 0.24(42) | **0.00(9)** | - | - | - |
| Hygiene | 0.00(4) | 0.00(2) | **0.00(1)** | - | - | - | 0.00(14) | 0.32(8) | **0.00(5)** | 0.61(108) | 0.52(55) | **0.00(10)** | - | - | - |
| Leisure | 0.00(15) | 0.00(8) | **0.64(2)** | 0.49(148) | 0.12(76) | **0.14(17)** | 0.54(98) | 0.53(49) | **0.48(10)** | 0.20(128) | 0.34(65) | **0.27(14)** | 0.18(88) | 0.10(44) | **0.29(8)** |
| Sleep | 0.94(265) | 0.94(134) | **0.98(2)** | 0.89(202) | 0.77(103) | **0.90(2)** | 0.93(198) | 0.96(100) | **0.83(19)** | 0.50(202) | 0.53(101) | **0.59(19)** | 0.94(172) | 0.90(87) | **0.97(16)** |
| Toileting | - | - | - | 0.20(8) | 0.18(6) | **0.60(6)** | 0.04(3) | 0.20(2) | **0.33(2)** | 0.00(21) | 0.11(14) | **0.00(4)** | 1.00(84) | 0.67(2) | **0.00(2)** |

<sup>†</sup>MoE: room-specialized Mixture-of-Experts model. <sup>‡</sup>Global Transformer: single monolithic baseline model.
Each cell reports the per-class F1 score, with the number of test segments (support) in parentheses. Within each site, the three columns correspond to window lengths $L = 5$ (regular), 10 (italic), and 60 (bold) minutes.

## IV. Discussion and Limitations

### A. The MoE-versus-monolithic-model gap

The proposed room-specialized MoE consistently outperformed monolithic baselines across all homes and window lengths. Rather than undersampling, the proposed approach decomposes ADL recognition into room-specific activity spaces. This reduces direct competition between dominant activities such as Sleep Rest and lower-frequency activities including Cooking, Dressing, and Medication. A likely explanation for the weaker baseline performance is that globally trained models allocate most representational capacity toward dominant classes in low-data residential settings, resulting in poor macro-F1 despite reasonable micro-accuracy. Because both the Global Transformer and the MoE experts rely on the same transformer-based sequence modeling framework, the observed performance differences are unlikely to arise from the underlying architecture itself. Instead, the results suggest that the primary advantage stems from the room-specialized decomposition strategy, which reduces class competition and allows each expert to focus on a smaller activity space. This finding is particularly relevant in residential sensing environments where data are limited, class distributions are highly imbalanced, and activities often occur within predictable spatial contexts[16,17]. Although the Global Transformer occasionally achieved relatively high micro-accuracy, its macro-F1 remained substantially lower. This divergence indicates strong bias toward dominant activities such as Sleep Rest and Leisure Social, inflating overall accuracy while masking poor recognition of minority ADLs.

### B. Effect of window length

Table 3 suggests that apparent performance gains at longer window lengths should be interpreted cautiously[18]. As temporal aggregation increases, the number of evaluation segments available for lower-frequency ADLs decreases substantially. In several cases, activities represented by only one or two test segments achieved perfect F1 scores, reflecting limited evaluation opportunities rather than robust discriminative performance. Consequently, improvements observed at longer window lengths may partly result from reduced sample counts rather than genuine gains in activity recognition capability. In contrast, shorter windows preserve a larger number of activity instances, particularly for infrequent ADLs such as cooking, dressing, and toileting, preserving a more representative evaluation set for infrequent ADLs such as cooking, dressing, and toileting.

### C. Personalized modeling and cross-home variability

Macro-F1 varied substantially across homes, ranging from 0.60 to 0.89. This variability reflects differences in home layouts, sensor placement, resident routines, and activity complexity: some room experts distinguished only two dominant activities, whereas others were required to separate multiple ADLs with highly similar sensor signatures. Table. 3 further shows that the activity composition is not uniform across sites, as some activities are unique to individual residents and their daily routines. This heterogeneity has a direct consequence for real-world deployment: a single pooled dataset cannot adequately represent the behavioral diversity across homes, which motivated our decision to model each home individually. These findings suggest that home-specific modeling is more appropriate than globally trained architectures for realworld residential sensing, and that optimal window length, routing structure, and expert configuration may differ across homes because each residential environment presents a distinct behavioral and sensing context.

### D. Limitations

This study has several limitations. First, the pipeline relied on caregiver-assisted retrospective ADL logs, which may introduce labeling noise and temporal boundary ambiguity despite the use of structured templates, standardized visual and electronic guidelines, predefined reporting schedules, and quality-control procedures tied to participant compensation. Second, recognition difficulty varied across rooms because some environments contained more overlapping activities with similar sensor signatures; in addition, the current sensor configuration could not directly distinguish participant activity from caregiver activity within shared spaces, limiting resident-level disambiguation. Third, expert assignment was derived from activity-to-room relationships observed in the annotated training data. While this deterministic routing strategy improves interpretability and reduces model complexity, future work should investigate adaptive gating mechanisms that can learn expert assignment directly from sensor observations. Finally, the study evaluated a relatively small number of homes, limiting generalizability across broader residential and behavioral settings. Obtaining longitudinal ADL annotations in real residential environments remains operationally challenging, particularly in cognitively impaired populations requiring caregiver-assisted logging. Future work will evaluate larger longitudinal deployments and incorporate presence-aware sensing mechanisms to improve resident-level activity disambiguation.

## V. Conclusion

We presented a deterministic-gated room-specialized MoE framework for in-home ADL recognition using ambient sensor data collected from community-dwelling older adults with early-stage dementia. Across five homes, the proposed MoE consistently outperformed globally trained transformer, CNN, and Random Forest baselines while using substantially fewer trainable parameters and maintaining efficient edge-device deployment on a Raspberry Pi 4. The findings suggest that architectural decomposition aligned with the spatial organization of residential ADLs may provide a practical and effective strategy for low-data, highly imbalanced ambient sensing environments. More broadly, the results support the importance of personalized, home-specific modeling approaches for realworld longitudinal monitoring of functional behavior in aging populations.

## Data Availability

All data produced in the present study are available upon reasonable request to the authors

## VI. Acknowledgment

This work is supported by Alzheimer’s Association Research Grant (24AARGD-NTF-1242722). The authors also like to acknowledge Sevara Pulatova, Abby Roessler, Claire Stam for their contribution to data collection.

